# Comparison of SARS-CoV-2 serological assays for use in epidemiological surveillance in Scotland

**DOI:** 10.1101/2021.04.20.21255596

**Authors:** Lindsay McDonald, Helen Wise, Frauke Muecksch, Daniel Poston, Sally Mavin, Kate Templeton, Elizabeth Furrie, Claire Richardson, Jaqueline McGuire, Lisa Jarvis, Kristen Malloy, Andrew McAuley, Norah Palmateer, Elizabeth Dickson, Theodora Hatziioannou, Paul Bieniasz, Sara Jenks

## Abstract

**Background:** Sero-surveillance of SARS-CoV-2 is crucial to monitoring levels of population exposure and informing public health responses, but may be influenced by variability in performance between available assays.

**Methods:** Five commercial immunoassays and a neutralising activity assay were used to detect antibodies to SARS-CoV-2 in routine primary care and paediatric samples collected during the first wave of the pandemic in NHS Lothian, Scotland as part of ongoing surveillance efforts. For each assay, sensitivity and specificity was calculated relative to consensus results and neutralising activity. Quantitative correlation was performed between serological and neutralising titres.

**Results:** Seroprevalence ranged from 3.4-7.3 % in primary care patients and 3-5.9 % in paediatric patients according to different immunoassays. Neutralising activity was detectable in 2.8 % and 1.3 % respectively. Relative assay performance changed depending on comparison to immunoassay consensus versus neutralising activity and qualititative versus quantitative agreement. Cross-reactivity with endemic seasonal coronaviruses was confirmed by neutralising assay in false positives for one immunoassay. Presence of false positives for another assay was found specifically in paediatric but not adult samples.

**Conclusions:** Five serological assays show variable accuracy when applied to the general population, impacting seroprevalence estimates. Assay performance may also vary in detection of protective neutralising antibody levels. These aspects should be considered in assay selection and interpretation in epidemiological studies.

## Introduction

The emergence of severe acute respiratory syndrome coronavirus 2 (SARS-CoV-2), the causative agent of COVID-19, represents a major global public health threat which has led to high excess mortality worldwide. Reverse transcription polymerase chain reaction (RT-PCR) detection of viral RNA in nasopharyngeal swabs serves as the standard method for diagnosis but is predicted to underestimate the burden of infection due to its narrow time window of detection and restriction of eligibility to symptomatic individuals. In Scotland, this underestimation was particularly pronounced for much of the first wave, when testing was only available for hospitalised patients and healthcare workers and their household contacts [1].

By contrast, serological assays test for previous infection and are therefore useful for population-based sero-epidemiological surveillance. These studies are key to informing public health response by estimating the true extent of exposure and potentially susceptibility within the overall population as well as specific demographic groups over the course of the pandemic. Increasing numbers of SARS-CoV-2 seroprevalence studies being reported from different parts of the world indicate high levels of variation between populations [2,3]. However, reliability of these findings may be significantly impacted by the variability in performance between different serological assays [4–6].

Over the course of a few months, commercial manufacturers have developed a vast number of immunoassays for detection of SARS-CoV-2 antibodies [7]. These are typically directed towards one of two immunogenic viral targets – nucleocapsid (N) and spike (S) proteins. As S protein facilitates host cell entry via its receptor binding domain, anti-S antibodies are therefore predicted to be neutralising [8,9]. To determine neutralising ability, which offers some indication of protective immunity, pseudotyped virus particle assays expressing SARS-CoV-2 S protein and a luciferase reporter have been shown to correlate well with neutralisation of authentic SARS-CoV-2 [10]. Variable correlation of neutralising titres with quantitative titres from different immunoassays has been shown in COVID-19 convalescent patients [11].

To monitor seroprevalence in Scotland, approximately 500 residual blood samples from primary care have been collected weekly since 20^th^ April 2020 [12]. An age/sex/geographical sampling frame is used to achieve a set representative of population demographics. The presence of SARS-CoV-2 antibodies is determined using a spike-targeted IgG assay from DiaSorin. In addition, paediatric samples from multiple sources were gathered between May and August to evaluate seroprevalence in children. Rates of exposure are particularly opaque among children, who are more likely than adults to experience asymptomatic or mild disease [13,14].

Initial Scottish evaluations of serological assay performance used pre-pandemic samples and serum from laboratory-confirmed COVID-positive patients collected ≥ 14 days post-PCR as negatives and positives respectively [15]. As a result, it may not accurately reflect assay performance in the overall population with a more heterogeneous spectrum of disease represented. It is therefore important to evaluate accuracy in a context specific to the intended use of epidemiological surveillance. Here, we compare performance of 5 immunoassays and a neutralisation assay for determining seroprevalence in adult and paediatric Scottish populations.

## Methods

### Samples

Primary care samples were selected from those collected in NHS Lothian for ongoing SARS-CoV-2 sero-surveillance. A total of 518 residual blood samples, collected for other diagnostic purposes in primary care settings, have been obtained from regional biochemistry laboratories across Scotland each week since 20^th^ April 2020. Sample numbers are stratified with equal numbers of male and female specimens from each of the following age groups - ≤ 20, 21-40, 41-64, ≥65. Our study used 355 samples collected in NHS Lothian from 3 weeks – w/b 27th April (118), 8th June (125) and 3^rd^ August (112).

237 paediatric samples were collected at point of discard from a mixture of hospital inpatients and GP patients between May and August. The set comprised 39 children aged 0-5, 44 aged 6-10 and 154 aged 11-18.

### Automated serological assays

Five commercial immunoassays designed for use in automated high-throughput analysers were used for the detection of antibodies to SARS-CoV-2: Abbott Architect SARS-CoV-2 IgG (anti-N), Roche Anti-SARS-CoV-2 total antibody (anti-N), DiaSorin LIAISON^®^ S1/S2 SARS-CoV-2 IgG assay, Siemens SARS-CoV-2 total antibody (anti-S), and EUROIMMUN Anti-SARS-CoV-2 ELISA (anti-S IgG). All generate a qualitative positive/negative result based on manufacturer-defined thresholds. Assays were performed on the Abbott Architect (NHS Lothian), Roche Elecsys (NHS Lanarkshire) DiaSorin LIAISON^®^ (NHS Lothian), Siemens Atellica (NHS Tayside) and EUROIMMUN Anti-SARS-CoV-2 ELISA (IgG)(Scottish National Blood Transfusion Service (SNBTS)) platforms in accordance with the manufacturers’ instructions.

### Neutralisation assays

Neutralising activity was measured using HIV-1_NL_ΔEnv-NanoLuc/SARS-CoV-2 pseudotype virus and seasonal coronaviruses HCoV-OC43, HCoV-NL63, or HCoV-229E as previously described [10,16]. Sera were initially diluted 1:12.5 then five-fold serially diluted in 96-well plates over four dilutions. Thereafter, approximately 5×10^3^ infectious units of the relavant virus were mixed with the serum dilutions at a 1:1 ratio and incubated for 1 hour at 37 °C. Mixtures were then added to 293T/ACE2cl.22 target cells (for HIV-1_NL_ΔEnv-NanoLuc/SARS-CoV-2 and HCoV-OC43) or HT1080/ACE2cl.14 (for HCoV-NL63 and HCoV-229E) plated the previous day at 1×10^4^ cells/well in 100 µl medium, giving a final starting serum dilution of 1:50. Cells were cultured for 48h and harvested for NanoLuc luciferase assay (SARS-CoV-2) or 24h and harvested for flow cytometry (OC43/NL63/229E). A half-maximal neutralisation titre (NT_50_) was calculated, with a limit of detection of 30 and positive/negative threshold of 50.

### Statistical analysis

All cohort samples (GP or paediatric as relevant) were included in seroprevalence and assay performance calculations. Neutralising activity was assumed to be negative in samples negative according to all immunoassays tested. For 4 immunoassay-positive samples which were insufficient (2 GP and 2 paediatric) neutralising activity was inferred based on findings for samples with similar serological results. Exclusion of these would have artificially skewed calculations due to the relatively low number of positive samples.

Spearman’s r correlations were generated using GraphPad Prism 9 to compare immunoassay and neutralisation titres for 28 primary care samples producing a positive result by any assay.

## Results

355 serum samples collected over 3 weeks in April, June and August were selected from a cohort of age and sex stratified primary care samples obtained from NHS Lothian biochemistry laboratories as part of ongoing sero-surveillance. These were analysed by 5 SARS-CoV-2 automated immunoassays. Seroprevalence was found to vary from 3.4-7.3 % depending on the assay used (Figure 1). Roche and Siemens assays produced concordant seroprevalence of 4.5 %. The DiaSorin and Euroimmun assays both identified unique positives – 11 and 3 respectively – resulting in greater seroprevalence. Discordant negatives led to the lowest seroprevalence (3.4 %) using the Abbott assay.

**Figure 1.**
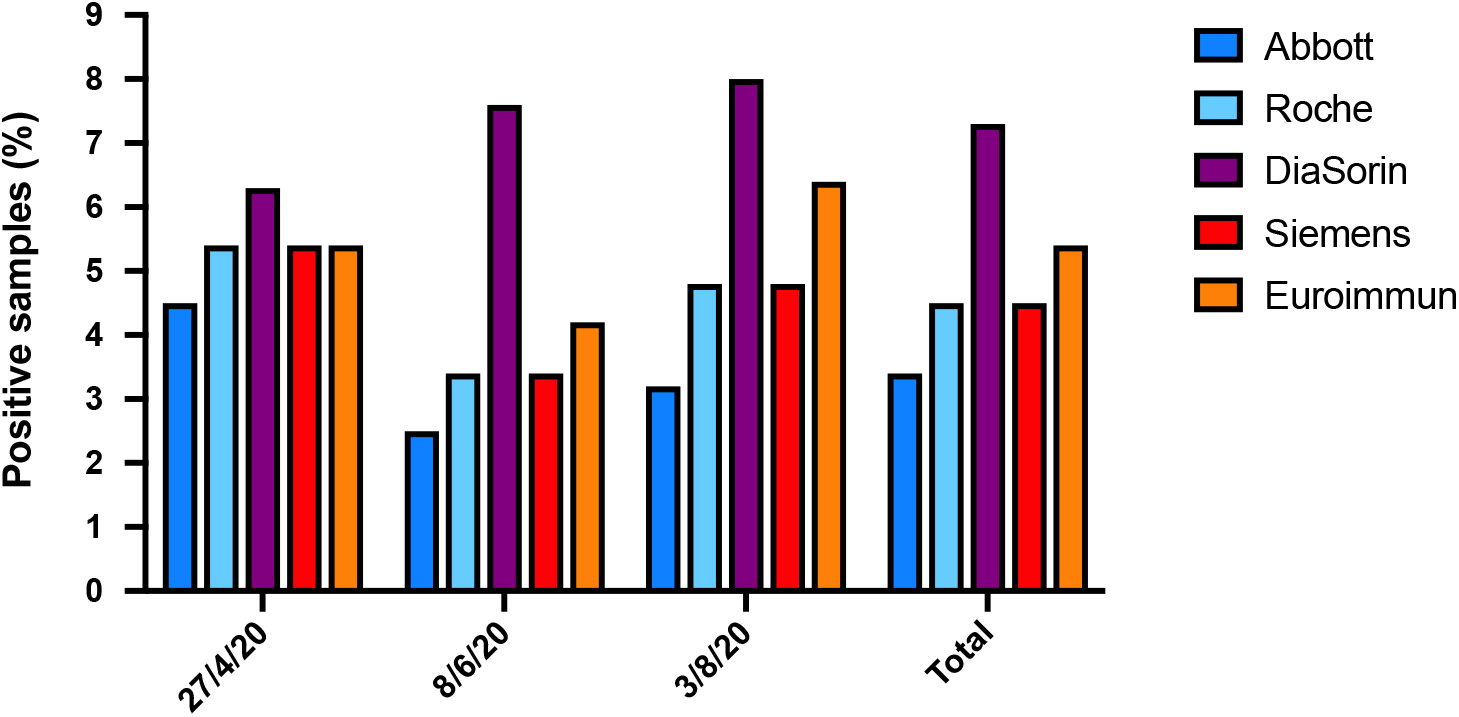
Detection of SARS-CoV-2 antibodies in 355 primary care samples.

To assess relative assay performance, true exposure status for each sample was inferred from consensus result (≥3/5 pos = pos), producing an overall seroprevalence of 4.5 %. Roche and especially Siemens assays performed well relative to this standard (Table 1). The Abbott assay, for which deteriorating sensitivity with time from infection has previously been described [11], showed the lowest sensitivity at 75 %. False positive results for DiaSorin and to a lesser extent Euroimmun in the context of relatively low population prevalence led to poor positive predictive values of 57.7 % and 84.2 % for these assays respectively.

**Table 1.** Comparison of assay performance relative to inferred exposure status. 355 primary care samples were tested for presence of SARS CoV 2 antibodies by 5 immunoassays. Exposure status was considered positive for samples found positive by 3 or more assays. Prevalence = 4.5 %.

|  | Abbott (N) |  | Roche (N) |  | Diasorin (S) |  | Siemens (S) |  | Euroimmun (S) |  |
| --- | --- | --- | --- | --- | --- | --- | --- | --- | --- | --- |
| No pos | 12 |  | 16 |  | 26 |  | 16 |  | 19 |  |
| % pos | 3.4 |  | 4.5 |  | 7.3 |  | 4.5 |  | 5.4 |  |
| FN | 4 |  | 1 |  | 1 |  | 0 |  | 0 |  |
| FP | 0 |  | 1 |  | 11 |  | 0 |  | 3 |  |
|  | % | 95 % CI | % | 95 % CI | % | 95 % CI | % | 95 % CI | % | 95 % CI |
| Sens | 75 | 47.6 to 92.7 | 93.8 | 69.8 to 99.8 | 93.8 | 69.8 to 99.8 | 100 | 79.4 to 100 | 100 | 79.4 to 100 |
| Spec | 100 | 98.9 to 100 | 99.7 | 98.4 to 100 | 96.8 | 94.3 to 98.4 | 100 | 98.9 to 100 | 99.1 | 97.4 to 99.8 |
| PPV | 100 | - | 93.8 | 67.9 to 99.1 | 57.7 | 42.9 to 71.2 | 100 | - | 84.2 | 63.4 to 94.3 |
| NPV | 98.83 | 97.3 to 99.5 | 99.7 | 98.1 to 100 | 99.7 | 98 to 100 | 100 | - | 100 | - |

To assess how prevalence of SARS-CoV-2 antibody related to presence of neutralising antibody, neutralisation activity was measured using a SARS-CoV-2 spike-pseudotyped HIV-1 virion expressing a nanoluc luciferase reporter. Neutralising antibody prevalence (2.8 %) was lower than for immunoassay. Neutralisation activity was not detected for any of the DiaSorin or Euroimmun single-positive samples.

Comparison of quantitative data showed that values from all 5 serological assays correlated with neutralisation titres to varying degrees (Figure 2). In general, relative strength of prediction of neutralising activity differed to that of qualitative results as indicated by PPV (Table 2). The Euroimmun assay came second in terms of titre correlation but fourth in terms of PPV, while the Roche assay came third in terms of PPV but was the poorest correlate.

**Table 2.** Comparison of immunoassay performance relative to neutralising activity positivity among primary care patients (n=355). Neutralising antibody prevalence = 2.8 %.

|  |  | Abbott |  | Roche |  | Diasorin |  | Siemens |  | Euroimmun |  |
| --- | --- | --- | --- | --- | --- | --- | --- | --- | --- | --- | --- |
| TP | FP | 9 | 3 | 9 | 7 | 10 | 16 | 10 | 6 | 10 | 9 |
| TN | FN | 342 | 1 | 338 | 1 | 329 | 0 | 339 | 0 | 336 | 0 |
|  |  | % | 95 % CI | % | 95 % CI | % | 95 % CI | % | 95 % CI | % | 95 % CI |
| Sens |  | 90 | 55.5 - 99.8 | 90 | 55.5 - 99.8 | 100 | 69.2 - 100 | 100 | 69.2 - 100 | 100 | 69.2 - 100 |
| Spec |  | 99.1 | 97.5 - 99.8 | 98 | 95.9 - 99.2 | 95.4 | 92.6 - 97.3 | 98.3 | 96.3 - 99.4 | 97.4 | 95.1 - 98.8 |
| PPV |  | 75 | 48.8 - 90.4 | 56.3 | 37.5 - 73.4 | 38.5 | 27.9 - 50.2 | 62.5 | 43 - 78.7 | 52.6 | 36.8 - 67.9 |
| NPV |  | 99.7 | 98.2 - 100 | 99.7 | 98.1 - 100 | 100 | - | 100 | - | 100 | - |

**Figure 2.**
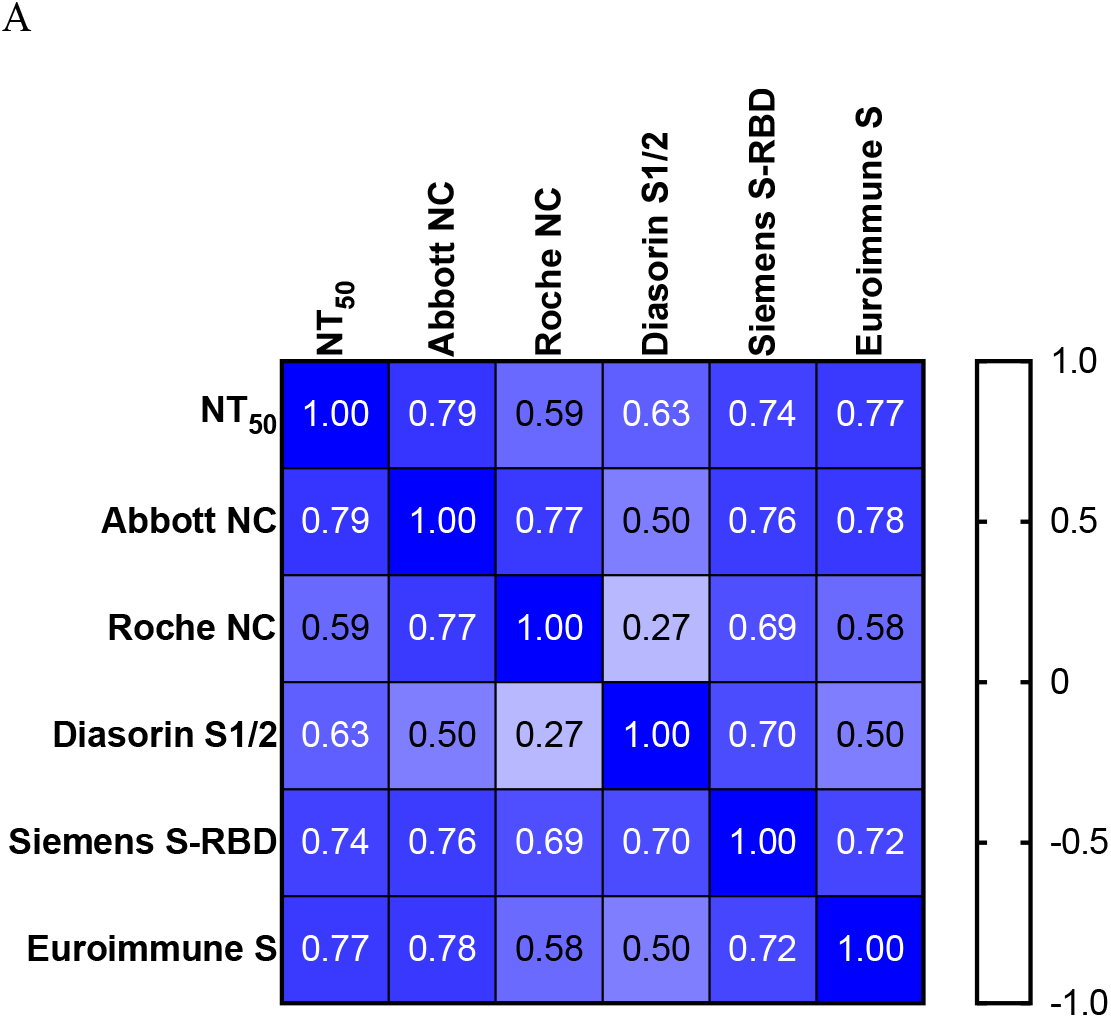
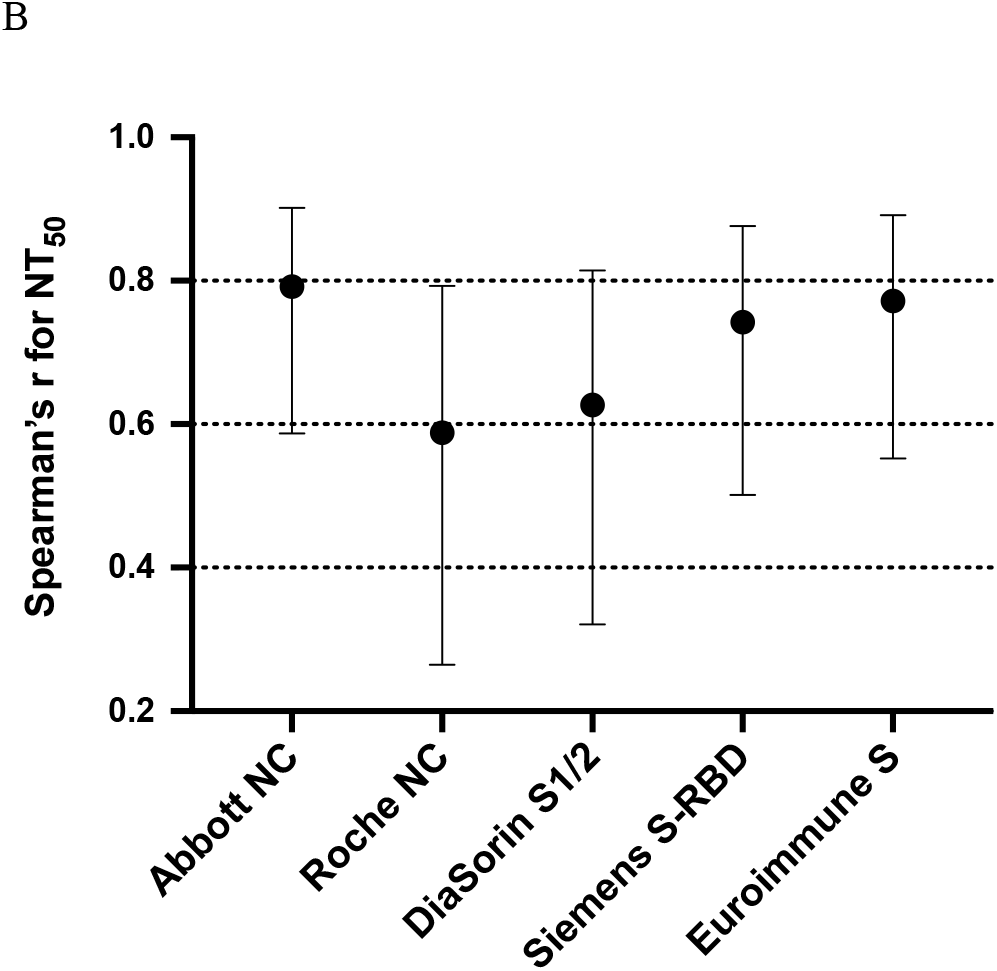
Correlation of serological assay titres with neutralisation titres in Ab-positive samples. A) Heatmap of Spearman’s r values for all comparisons. B) Spearman’s r values of serological assays to NT_50_ showing confidence intervals. NT_50_ LOD=30.

Prevalence was investigated in children using 237 paediatric samples collected from hospital inpatients and primary care patients between May and August. Samples were analysed by Roche, DiaSorin and Siemens immunoassays and a consensus positive result assigned to samples positive by all 3. All other positive results were according to a single assay only. Consensus results gave an exposure rate of 3 % (Figure 3). Neutralising activity was only detected in 3 samples, giving a very low prevalence of 1.3 %. All 3 immunoassays were poor predictors of neutralising activity positivity owing to its low levels within this cohort (Table 3). Interestingly, while Roche results agreed with the immunoassay consensus, the Siemens assay produced a number of unique positive results, which was not observed in the adult set. These samples did not display neutralising activity and are most likely false positives, indicating the presence of Siemens cross-reactive antibody or other component potentially specific to children.

**Table 3.**
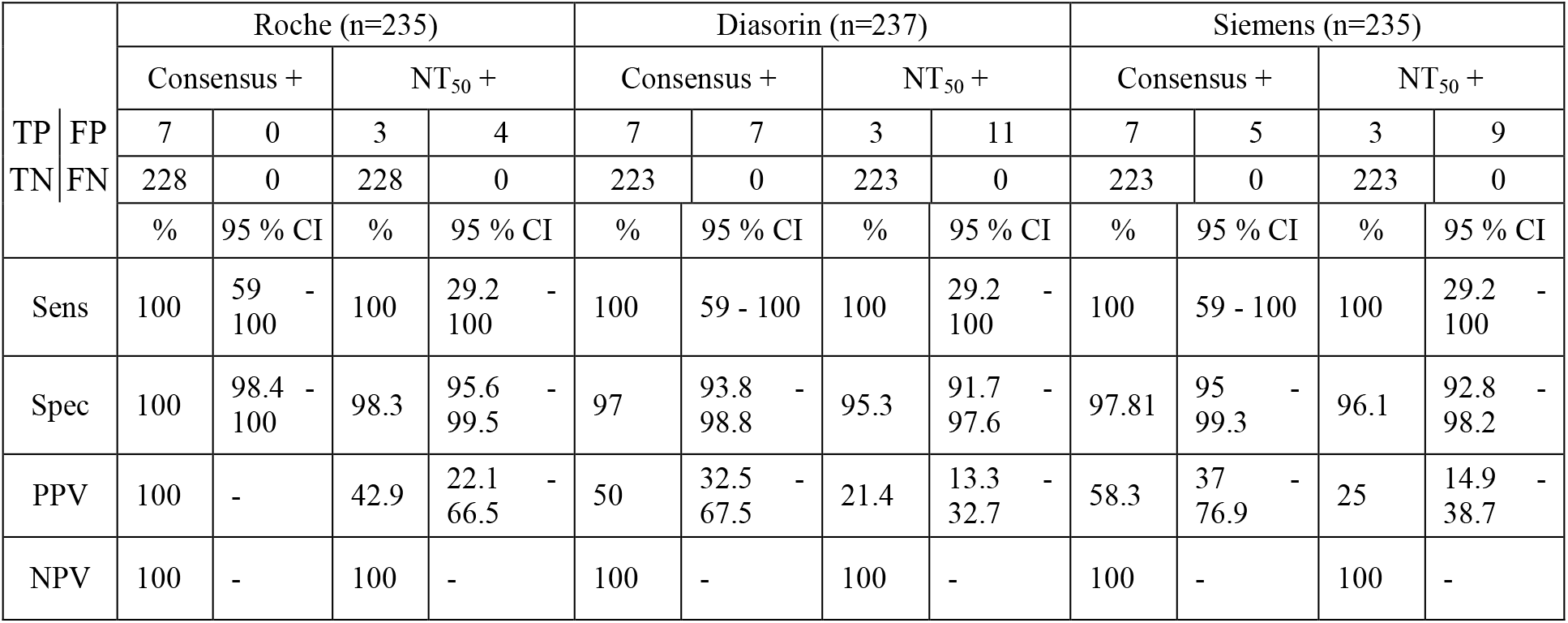
Comparison of assay performance in children relative to inferred exposure status (consensus 3/3+; 3 % prevalence) and neutralising antibody positivity (NT_50_+; 1.3 % prevalence).

**Figure 3.**
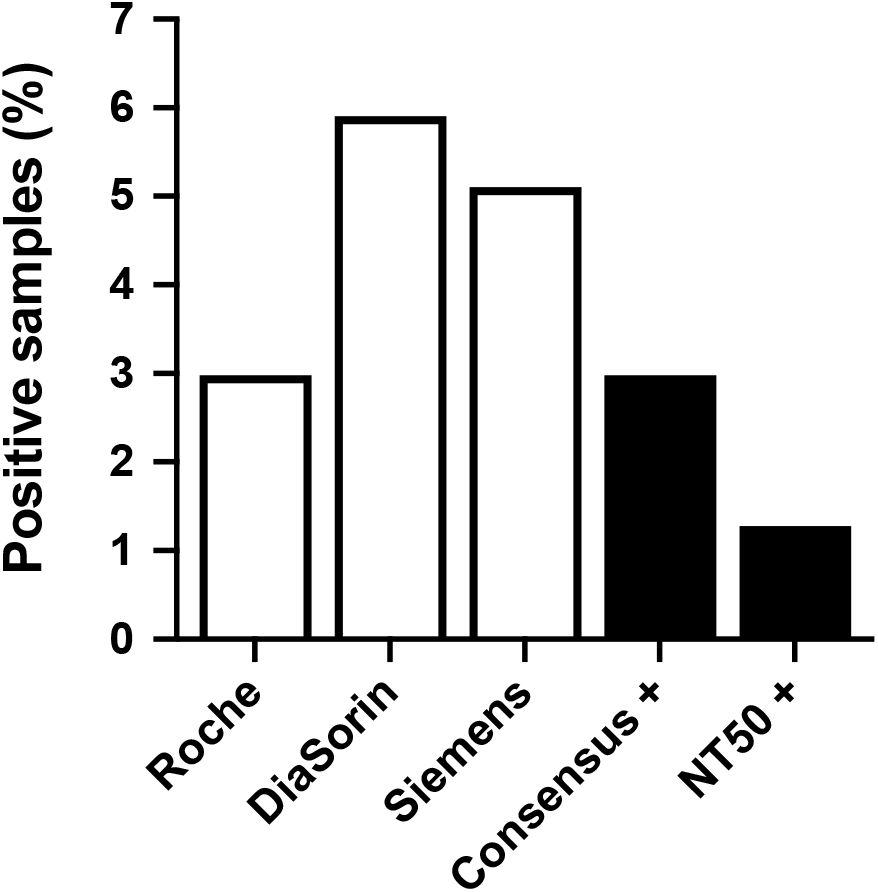
Seroprevalence of SARS-CoV-2 antibody in paediatric samples according to immunoassays and neutralising activity assay. Consensus+ = positive for 3/3 immunoassays.

One obstacle to specificity of SARS-CoV-2 immunoassays is the presence of antibodies to other human coronaviruses within the population. To investigate the possibility of cross-reactivity, 5 DiaSorin-only positive paediatric samples were tested for neutralising activity against endemic seasonal coronaviruses OC43, NL63 and 229E. Four out of five were found to be positive for at least two of the seasonal coronaviruses tested. None showed neutralising activity against SARS-CoV-2. This indicates that the DiaSorin assay produces false positives as a result of cross-reactivity with seasonal coronaviruses which does not reflect cross-protection *in vivo*.

## Discussion

Sero-epidemiological surveillance can provide information about population exposure and infection dynamics crucial to the public health response to the COVID-19 pandemic, but outcomes are influenced by choice of serological test. We found that over a subset of weeks from different stages of the first wave in Lothian, seroprevalence levels ranged from 3.4-7.3 % in primary care patients and 3-5.9 % in paediatric patients depending on the immunoassay used.

The use of biochemistry samples collected for other diagnostic purposes provides ready availability of samples from a broad spectrum of the population for sero-surveillance testing. Admittedly, the paediatric cohort may be less typical of Scottish children due to the inclusion of emergency admission samples, complicating direct comparisons with the mostly adult primary care dataset. However, our identification of lower seroprevalence in children – 3 % vs 4.5 % in the mostly adult primary care cohort – is consistent with studies from other parts of the world [17–20]. SARS-CoV-2 has been shown to be less efficient in infecting children than adults [21–23],. However, equivalent susceptibility in children to adults has been postulated as an explanation for the increased transmission dynamics observed with the new B.1.1.7 variant associated with the most recent wave of infections in the UK [25]. If this is the case, it may be observed that paediatric seroprevalence trends upwards towards that of the general population in coming months.

The DiaSorin assay is currently the primary tool for ongoing serological surveillance in Scotland [12]. In this assay comparison it showed the poorest agreement with consensus antibody positivity in both GP and paediatric sets. Specificity is known to be lower than for other assays investigated here [15,26], and a correction factor is accordingly applied [12] to enable a more accurate estimation of true sero-prevalenceNeutralisation assays demonstrated that false positives are generated due to assay cross-reactivity with antibodies to endemic seasonal coronaviruses. These antibodies are not cross-protective, as no neutralising activity against SARS-CoV-2 was detectable. Interestingly, boosting of seasonal coronavirus antibodies in response to SARS-CoV-2 infection with no associated protective effect has recently been described, indicating that such antibodies may also be cross-reactive *in vivo*, if non-neutralising [27].

The Euroimmun assay was also subject to false positives and consequently low PPV, although to a lesser degree than DiaSorin. This assay is currently used by the SNBTS and other UK transfusion centres to measure antibody levels in convalescent plasma donors. However, quantitative titres are used for this purpose, and its selection is based on demonstration of optimal correlation to neutralisation titres relative to alternative commercial and non-commercial immunoassays [28,29]. In accordance, our data confirmed a strong correlation between Euroimmun and neutralisation assay titres.

The Abbott assay demonstrated the lowest sensitivity relative to the consensus. It has previously been demonstrated that the sensitivity of this assay declines sharply with increasing time from infection, an effect not observed in Roche, DiaSorin or Siemens assays. This may account for it showing the highest level of correlation to neutralising titre, which has also been shown to decrease with time [11]. However, overlap in 95% confidence intervals for the Spearman’s R correlation coefficients indicates that it is not possible to conclude significantly improved correlation with any one assay. It has also been shown in previous studies that there is no correlation between Abbott titre and NT_50_ at the individual patient level [11]. Its utility in estimating previous infection is therefore liable to diminish as we move farther from the first wave. This was highlighted by the SNBTS’s finding that seroprevalence increased from 1.4 % by Abbott measurement to 5.4 % using Euroimmun in a one week comparison in October 2020 (unpublished data).

Optimal performance was achieved using the Roche N assay across both sample sets, in agreement with high sensitivity and specificity identified in evaluations performed in known positive and negative cohorts [15,26,30]. While this recommends its use in estimation of previous infection, poor quantitative correlation with neutralisation titre suggests it is less optimal as an indicator of immunity. Intriguingly, while the Siemens assay achieved perfect specificity in the GP sample set, it identified 5 discordant positives in the paediatric set. These samples were negative by other immunoassays and showed no neutralising activity and can be reasonably presumed to be false positives. Their exclusivity to paediatric samples raises the possibility of cross-reactivity with antibodies to a childhood virus or component otherwise specific to or primarily affecting children, and highlights the importance of evaluating assay performance in different population groups.

While our immunoassay findings can offer a reasonable estimate of previous exposure to SARS-CoV-2, this does not necessarily correspond to protection from subsequent infection. Neutralising antibody is undetectable in a proportion of convalescent patients [31,32] and seroprevalence estimates are consequently lower using neutralisation tests than immunoassay methods [2], a finding confirmed by our study. Although unlikely, the presence of neutralisation activity in additional seronegative samples, which were not tested, cannot be excluded. We found substantial rearrangement of relative assay performance when judged by quantitative correlation to neutralisation titre in place of consensus accuracy. This highlights the importance of targeting assay selection to specific use, as the optimal assay for estimating previous infection does not match that for assessing levels of protection.

One limitation of our study is that calculations are based on relatively small numbers of positive samples. Given the low population prevalence, larger cohorts would be required to conclusively determine optimal immunoassays for correlation with neutralising antibody activity. It is also worth noting that all samples included predate the emergence of epidemiologically significant new variants. Their impact on assay performance for seroprevalence therefore remains unknown.

This study demonstrates the dependence of SARS-CoV-2 assay suitability on population and study-specific factors. Ongoing investigation of these will be critical to reliable assessment of levels of exposure and protection in the face of evolving factors including waves of infection, new variants and population vaccination.

## Data Availability

Raw de-identified serological and NT50 data is available from the authors on request.

